# Knowledge-Driven Online Multimodal Automated Phenotyping System

**DOI:** 10.1101/2023.09.29.23296239

**Authors:** Xin Xiong, Sara Morini Sweet, Molei Liu, Chuan Hong, Clara-Lea Bonzel, Vidul Ayakulangara Panickan, Doudou Zhou, Linshanshan Wang, Lauren Costa, Yuk-Lam Ho, Alon Geva, Kenneth D. Mandl, Suchun Cheng, Zongqi Xia, Kelly Cho, J. Michael Gaziano, Katherine P. Liao, Tianxi Cai, Tianrun Cai

**Author notes:** Xiong, Sweet and Liu contributed equally. Cai and Cai contributed equally.

## Abstract

Though electronic health record (EHR) systems are a rich repository of clinical information with large potential, the use of EHR-based phenotyping algorithms is often hindered by inaccurate diagnostic records, the presence of many irrelevant features, and the requirement for a human-labeled training set. In this paper, we describe a knowledge-driven online multimodal automated phenotyping (KOMAP) system that i) generates a list of informative features by an online narrative and codified feature search engine (ONCE) and ii) enables the training of a multimodal phenotyping algorithm based on summary data. Powered by composite knowledge from multiple EHR sources, online article corpora, and a large language model, features selected by ONCE show high concordance with the state-of-the-art AI models (GPT4 and ChatGPT) and encourage large-scale phenotyping by providing a smaller but highly relevant feature set. Validation of the KOMAP system across four healthcare centers suggests that it can generate efficient phenotyping algorithms with robust performance. Compared to other methods requiring patient-level inputs and gold-standard labels, the fully online KOMAP provides a significant opportunity to enable multi-center collaboration.

## 1 Introduction

In healthcare institutions, electronic health records (EHR) are commonly utilized to electronically store and manage patients’ records, encompassing diverse data such as demographic details, diagnostic billing codes, medical procedures, laboratory test outcomes, medication prescriptions, and narrative notes collected by healthcare professionals during patient vis its. The extensive volume of EHR data, both in terms of sample size and data elements, presents a valuable resource for translational research. For instance, it can be utilized to identify cohorts of patients suitable for clinical studies (1), develop clinical decision support tools to optimize patient outcomes (2), and pinpoint possible biomarkers and therapeutic targets for various diseases. Additionally, EHR data can provide real-world evidence for comparative effectiveness studies involving a broader population than randomized clinical trials (3; 4), and by being linked with biobanks, can aid in genetic research on the origins of diseases (5). EHRs also enable researchers to quickly assemble retrospective cohorts of large sample sizes with a broad range of outcomes and predictors, making clinical research more efficient and cost-effective. Moreover, since EHR data is closely related to patient care, models trained on it can be more readily applied to clinical practice. Consequently, EHR-driven research is more efficient than traditional cohort studies and has the potential to expedite the translation of research outcomes into clinical practice.

Fully realizing the translational potential of EHR data, however, has proven to be a significant challenge in part due to the imperfect nature of EHR data. Taking research studies on disease progression or treatment response for rheumatoid arthritis (RA) patients as an example, the first step of such a study is often to assemble a study cohort consisting of RA patients. However, one major limitation of EHR data is that precise information on disease phenotype, such as the presence of RA, is not readily available (6). Diagnostic billing codes often do not accurately reflect the true disease status (1). This could be due to uncertainty in the diagnosis, especially during the early stage of the disease, as well as inconsistencies in coding practices. In an EHR study with data from Mass General Brigham (MGB), only 58% of those patients with at least 3 International Classification of Diseases (ICD) codes of RA were confirmed RA cases (7). Forming disease cohorts based on inaccurate ICD codes can lead to substantial biases in downstream EHR data analyses (8).

Recent efforts have been directed towards enhancing the accuracy of assigning disease phenotypes while maintaining scalability of EHR driven studies. This has led to the development of label-efficient, high-throughput, semi-supervised or weakly supervised machine learning algorithms that can precisely identify disease phenotypes (4; 9). Since relevant information pertaining to a phenotype is often dispersed across different fields in the EHR, including unstructured clinical notes, weakly supervised techniques that utilize multiple features have proven to be effective across diverse healthcare systems and disease phenotypes. (10; 11; 12; 13). Weakly supervised approaches typically rely on noisy labels, such as ICD codes and/or mentions of clinical terms representing the target disease in narrative notes, which can be extracted via natural language processing (NLP). These noisy labels are used to train algorithms that improve upon the disease ICD code or NLP mention alone by combining information from additional features that serve as supporting evidence. Including these supporting features is particularly important for disease phenotypes without highly accurate ICD codes (12).

Although weakly supervised approaches have shown potential, current algorithms rely on user input to define the feature set for a given phenotype, which could greatly restrict their scalability. Existing automated feature selection methods such as the SAFE (Surrogate assisted feature extraction) procedure (14) typically require patient level data.

Moreover, all existing phenotyping algorithms also require patient-level data for training and validation, which significantly curtails the ability of research teams from different institutions to collaborate. To overcome these challenges, we developed a knowledge-driven online multimodal automated phenotyping (KOMAP) system that generates a list of informative features and enables the training of a multimodal algorithm based on simple summary data provided by the user.

The KOMAP system includes an online narrative and codified feature search engine (ONCE) to assist researchers to generate a list of informative codified and narrative features that can be used as input features for predictive modeling of a target disease phenotype. The ONCE system is powered by knowledge graph embeddings trained using multi-source representation learning of EHR concepts. With ONCE selected features, KOMAP Web Ap plication Programming Interface (API) can train a phenotyping algorithm online based on a user-supplied summary of the feature matrix. When a small number of gold standard labels are available, KOMAP can also provide a highly reliable estimate of the algorithm performance based on additional summary statistics. Real world validation of the KOMAP system using EHR data from multiple healthcare centers suggest that it can generate phenotyping algorithms with robust performance. With online feature selection and algorithm training capability using summary level data, the KOMAP system has the potential to increase the translational value of EHR data by efficiently improving the resolution of EHR data and facilitate cross-institutional collaborations.

## 2 Methods

The KOMAP pipeline comprises two key components: (i) the online narrative and codified feature search engine (ONCE) powered by multi-source knowledge graph representation learning; and (ii) the online phenotyping algorithm training and validation.

### 2.1 ONCE Powered by Representation Learning

EHR features include both codified and narrative features. We focus on four broad categories of EHR codes mapped to common ontologies: (i) ICD codes are grouped and mapped to PheCodes (15); (ii) medication prescriptions are mapped to ingredient level RxNorm codes (16); (iii) all ICD and CPT procedure codes are grouped according to the Clinical Classifications Software (17); (iv) laboratory tests are mapped to LOINC codes (18). For narrative features, we extract mentions of clinical terms from free text clinical notes via the Narrative Information Linear Extraction (NILE) (19) software, which normalizes all clinical terms to standardized clinical concepts represented by concept unique identifiers (CUis) according to the unified medical language system (UMLS) (20). To identify an initial set of clinical concepts important to a target disease, the ONCE system additionally leverages a large knowledge base of medical articles that describe the relatedness between narrative features and the disease at a higher level. The current article corpus comprises data from seven online sources including Wikipedia, Medscape, and Merck Manuals. We apply NILE to the knowledge source articles to extract narrative features found in the article text as well as the title of each article.

For a target disease with its name as input, the ONCE system can generate a list of codified features and a list of narrative features. The key component that powers the ONCE feature selection engine is the semantic representations of all EHR codified and NLP concepts, trained via multi-source knowledge graph representation learning which we describe in Section 2.1.1. To further improve specificity of the selected NLP concepts, ONCE only selects CUis from a list of those that appear in knowledge source articles with titles relevant to the target disease as detailed in Section 2.1.2. The final ONCE selection criteria are based on a composite score that integrates information on the relatedness of a candidate concept to the target concept as quantified by the semantic embeddings, the frequency of the candidate concept in the EHR, and a weighted frequency of the concept in the knowledge source articles. Figure 1 presents a graphical illustration of ONCE.

**Figure 1:**
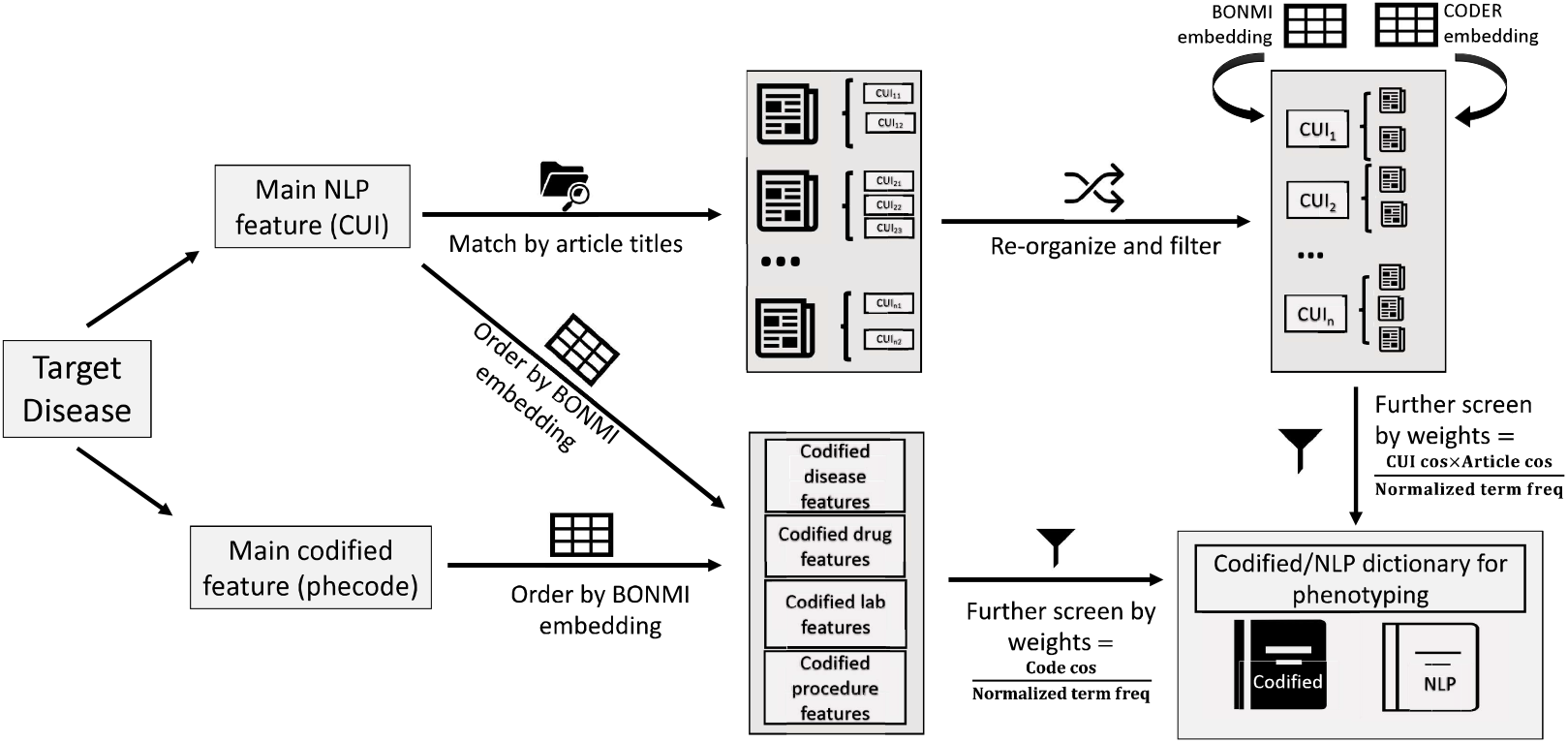
An illustration of how ONCE app generates codified and NLP phenotyping dictionaries for a user-specified disease.

#### 2.1.1 Multi-source Representation Learning (MultiReL) of EHR Concepts

To train a robust semantic representation of EHR concepts, we synthesized three sources of information: (i) Embeddings for 60,106 concepts pre-trained using a variant of the Skip-Gram algorithm with longitudinal EHR data from 12.5 million patients at the VA (21); (ii) pre trained embeddings for 84,995 concepts using Mass General Brigham Biobank (MGBB) EHR data with 60k patients; (iii) pre-trained contextual embeddings for the textual descriptions of the clinical concepts that appear in either online knowledge sources (including the article titles), or the two EHR systems via the large language model (LLM) based CODER algorithm (22). EHR concepts need to be present in both healthcare systems, or have a frequency of over 5% to be considered. To integrate the two sets of EHR embeddings from VA and MGB, which have overlapping but not identical concepts, we employ the Block-wise Overlapping Noisy Matrix Integration (BONMI) algorithm (23) to generate a fused EHR embedding for the union of the EHR concepts appearing in both institutions, denoted by 𝕍_EHR_· The input of the BONMI algorithm contains the two embeddings trained by data in VA and MGBB separately. (23) has shown improved performance due to the knowledge integration through BONMI, compared to the initial embeddings generated from data from a single institution.

To create the LLM-driven embeddings for the CUis, we first generate term-level embed dings via CODER, and then for each CUI, we create their LLM embeddings as the average of the embeddings across all terms that represent the CUI. CODER additionally creates embeddings for all article titles and codified features. Since the LLM embeddings and the EHR embeddings are trained in different context, we train an orthogonal translation matrix between the two sets of embeddings using CUis that are shared in the two sources and subsequently map all LLM embeddings to the EHR space, denoted by 𝕍_LLM→EHR_ which includes 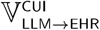 for individual NLP concepts, 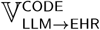 for individual codified concepts, and 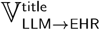 for article titles. The final MultiRel embeddings concatenate the BOMNI embeddings with the LLM trained embeddings 𝕍_LLM→EHR_ to leverage information from different representations. These MultiReL EHR concept embeddings power the ONCE search engine.

#### 2.1.2 ONCE Search Engine

For a user-specified disease name, the ONCE search engine starts by identifying online knowledge source articles that are relevant to the disease. For a given disease CUI, the relevance of articles will be ranked according to the cosine similarity between their 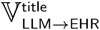 with 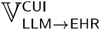 of the disease, denoted by cosine^target→title^. Only articles with titles attaining cosine^target→title^, exceeding a threshold, which is set to 0.5 by default, are selected as relevant for the target disease. We restrict the search to all clinical terms that can be mapped to a CUI in the UMLS and also appear in the selected knowledge source articles. Within the CUis appearing in the selected articles deemed relevant to the target disease, we further create a relevance score for each candidate CUI that combines multiple sources of information: (i) the cosine similarity between the candidate CUI and the target CUI based on the concatenated embeddings 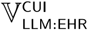 denoted by cosine^target→CUI^; (ii) a weighted frequency of the candidate CUI appearing in the knowledge source articles which reflects its impor tance according to literature denoted by 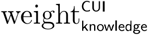; and (iii) the log frequency of the CUI appearing in the EHR denoted by 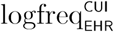. The knowledge source weight, 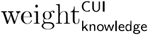, is calculated as the sum of cosine^target→title^ for all articles selected for the target disease and contain the CUL A CUI gets a higher weight when it appears in more articles and articles with titles less relevant to the CUI will be discounted in this step. To avoid the disproportionate contribution of some rare features’ frequency, we normalize 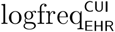 as 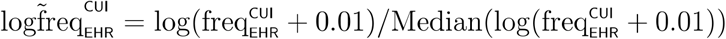, where the median is taken over relevant CUis. Combines the three sets of weights, ONCE assigns a final relevance score to a candidate CUI as

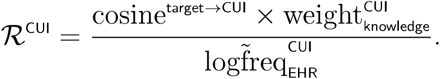

For a given target disease, ONCE assigns relevance scores for all CUis that appear in the selected articles and belong to EHR, denoted by Ω^target^. We further screen out less relevant features by evaluating the distribution of {log(ℛ^CUI^), CUI ∈ Ω^target^}. We further remove those CUis with log(ℛ^CUI^) lower than the mode plus median absolute deviation of {log(ℛ^CUI^), CUI ∈ Ω^target^}. Note that ONCE selected features can also be used to perform clinical studies on the disease rather than for phenotyping. We suggest a lower threshold value (e.g., only remove CUis with log(ℛ^CUI^) lower than the mode) for clinical studies, which focus on association detection and require a more comprehensive dictionary of related features for the disease.

The EHR embeddings also provide representations for codified features. Yet, directly mapping the unstructured text in the online knowledge sources to the codified clinical fea tures may introduce ambiguity and noise. Therefore, we identify important codified features based on a simplified form of ℛ^CUI^ as 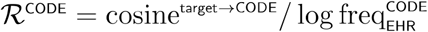, where cosine^target→CODE^ is based on 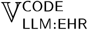. We then apply the same inclusion criteria that only codes with log(ℛ^CODE^) larger than the mode plus median absolute deviation of log(ℛ^CODE^) are kept.

### 2.2 KOMAP Algorithm Training and Validation

With a given set of selected features, including a set of main surrogate features and a healthcare utilization measure denoted byℋ, KOMAP algorithm can be trained online based on the covariance matrix of x ↦ log(x *+* 1) transformed counts of these features. Although ONCE serves as a convenient and powerful tool for feature selection, the KOMAP algorithm can use any set of input features, provided that they include at least one highly predictive surrogate feature such as the main PheCode or CUI representing the disease. The key steps of KOMAP training and validation are illustrated in Figure 2. Let **X** = (*X*_1_, …, *X*_*p*_)^⊤^ denote the set of input features; and let {**X**_*i*_, *i* = 1, …, *N*} denote the observed feature data on *N* subjects from a population targeted for the phenotyping algorithm. The empirical covariance matrix of **X**, denoted by ℂ, is the main input for the KOMAP algorithm. Without loss of generality, we assume that the first *K* features, *X*_1_, …, *XK*, are the available main surrogates; and *X*_*p*_ is the utilization measure ℋ.

**Figure 2:**
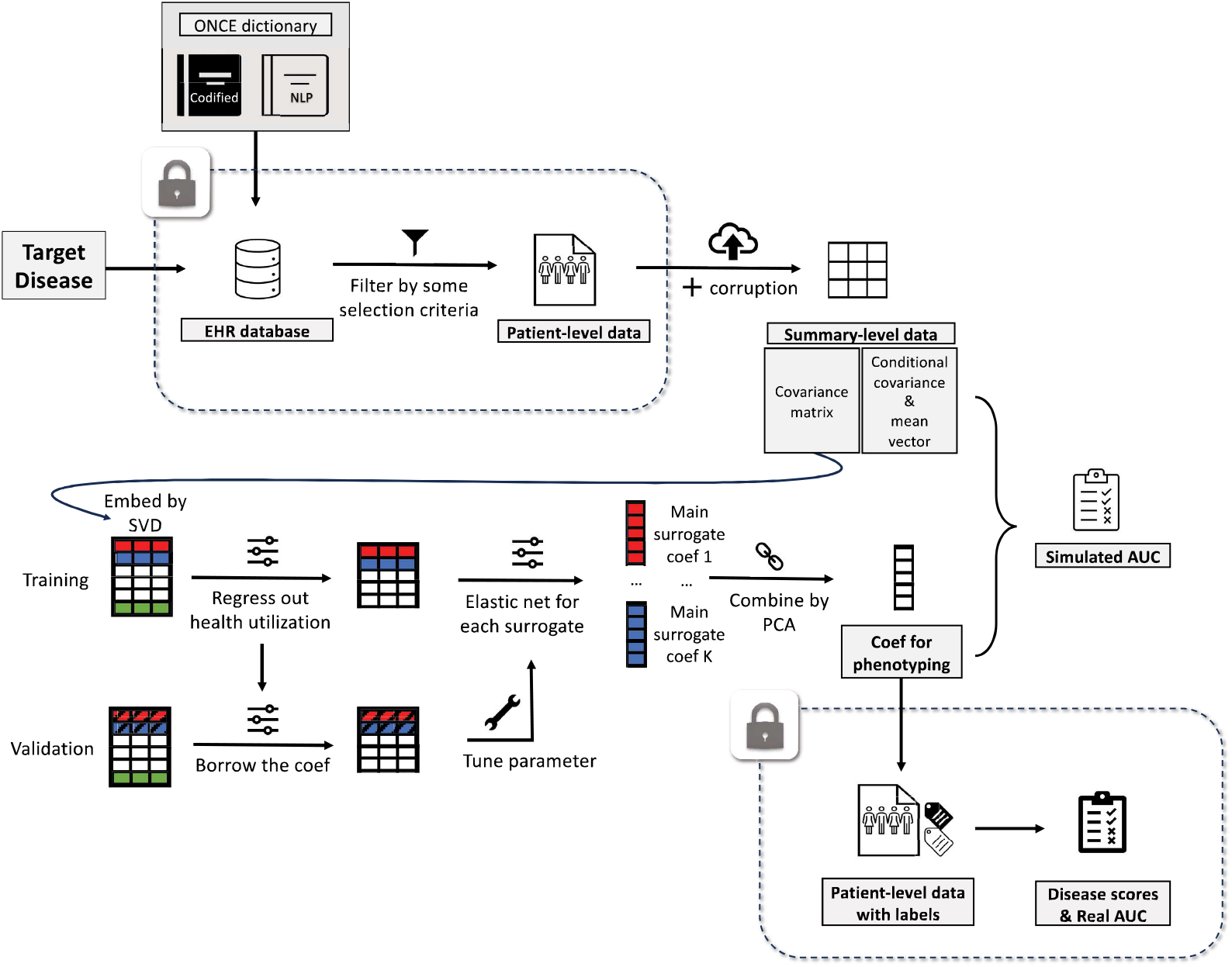
Flowchart of the KOMAP online phenotyping algorithm.

#### 2.2.1 KOMAP training

In this section, we shall introduce the training algorithm with its mathematical details in cluded in Supplementary Materials B.l. The algorithm contains three steps including nor malizing the main surrogates with the utilization, denoising via regression on each main sur rogate, and combining the derived risk scores of different surrogates, all of which only require the embedding of ℂ free of any individual-level information. With a given ℂ, KOMAP starts by performing the Cholesky decomposition ℂ _*p*×*p*_ = 𝕌 ^⊤^ 𝕌, where ℂ _*p*×*p*_= [𝕌_1,…,_ 𝕌_*K,…*,_ 𝕌_*P*_], 𝕌*i* represents embedding vectors of the *i*th feature with dimension *p*. We first remove the effect of *X*_*p*_ on each of the main surrogates by regressing 𝕌_*k*_ ∼ 𝕌_*p*_ to obtain residual 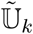 for *k* = 1, …, *K*, viewed as a way of normalization; and then reconstruct a new embedding matrix 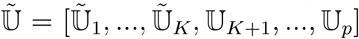. Subsequently, for *k* = 1, …, *K*, we regress 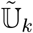 against 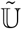 via an elastic net penalized linear regression (24):

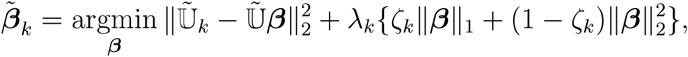

where λ_*k*_ and ζ_*k*_ are two tuning parameters. Then we derive the denoised embedding matrix for the main surrogates denoted as 𝕎 = (𝕎_1_, 𝕎_2_, …, 𝕎_*K*_) where 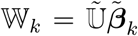. W can be viewed as a representation of the individual-level risk scores obtained using the same steps outlined above. At last, we combine the derived scores in 𝕎 through principle component analysis (PCA):

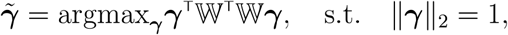

which aims at finding the most representative direction of the risk scores. The final phe notyping score of the embedding 𝕌 can be written as 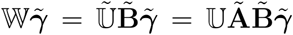, where the coefficient matrix 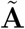 is constructed with the regression coefficients of 𝕌_*k*_ ∼ 𝕌_*p*_ obtained in our first normalization step; see Supplement B.1 for its formulation. Thus, the transferable coefficient is 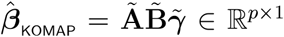 and our KOMAP training algorithm finally ranks the likelihood of patients having the disease according to 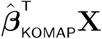.

Interestingly, this training algorithm is designed in such a way that all its specific steps, including adjustment against utilization, elastic net denoising, and PCA are based on linear models having the empirical covariance ℂ as the sufficient statistics for their implementation. Meanwhile, the embedding matrix 𝕌 shares the same covariance structure as the individual data matrix. Thus, 𝕌 can be used to produce *exactly* the same results obtained with the individual level data using our algorithm. We show this equivalence property in Supplement Materials B.l. The ridge component of the elastic net penalty mimics the dropout training as a regularization (25) and the lasso regularization achieves sparsity in the final trained algorithm. The choice of the tuning parameters λ_*k*_ and ζ_*k*_ is critical to the performance of the algorithm. We develop a data-driven tuning strategy that involves an additional covariance matrix constructed by sample splitting and data corruption as detailed in Supplement B.2.

### 2.2.2 KOMAP validation

When a small number of labels are available for validation, one may evaluate the performance of 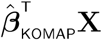 for classifying the disease status *Y* based on patient level data. Exact evaluation of the KOMAP algorithm with respect to performance metrics such as the receiver operating characteristic (ROC) and the area under the ROC curve (AUC) would require linking the results of the KOMAP algorithm to back to patient level data for the calculation, which would require high communication cost especially when multiple institutions are collaborating. We developed an online evaluation system to approximate such accuracy parameters based on additional summary statistics derived from a validation set consisting of labeled data 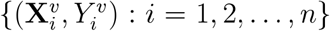, where the gold standard labels 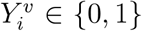 on the true disease status are typically ascertained through chart review. The number of labels should guarantee that the conditional sample mean and covariance of features are a good approximation of their population version. The key *working* assumption behind the proposed evaluation algorithm is that **X** I *Y* = *y* follows approximately *N*(***µ***_*y*_, **∑**_*y*_) for *y* ∈ {O, 1}, which implies that 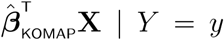is approximately 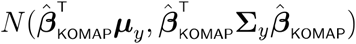. With this normal mixture assumption, the ROC curve of 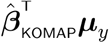 is uniquely determined by the mean vectors {***µ***_*y*_, *y* = 0, 1} and covariance matrices {**∑**_*y*_, *y* = 0, 1}. The accuracy parameters can be approximately calculated based on observations simulated from 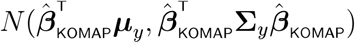. for *y* = 0, 1. See Supplementary Materials B.3 for more details. Unlike KOMAP training, this evaluation algorithm taking summary data as its input may not produce exactly the same results as that obtained using individual-level data. Nevertheless, since the gaussian mixture assumption tends to hold approximately for summary scores of EHR features (26, e.g.), especially under a high-dimensionality of *p* (27; 28), our KOMAP online validation algorithm can typically well approximate the true ROC and AUC as illustrated in real-world applications.

### 2.3 Validation Studies

#### 2.3.1 Evaluation of ONCE Embeddings and Feature Selection

We evaluated the quality of the EHR embeddings trained via MultiReL by assessing their ability in detecting known relationships between EHR concept pairs. We curated different categories of known relations, largely as including similarity and relatedness, from online knowledge sources including the UMLS and existing ontologies such as RxNorm and LOINC. Similar pairs of codified concepts were largely created based on code hierarchies including the PheCode hierarchy. Since a majority of laboratory codes in the VA are not mapped to LOINC codes, we augmented the LOINC hierarchy with manually annotated similar pairs when assessing similar laboratory code pairs. Similar CUI pairs are extracted from relationships in the UMLS. We additionally evaluated the similarity between mapped CUI ⟷ code pairs. We leveraged UMLS to obtain the mapping from different medical coding systems to concept unique identifiers (20) and performed semi-automated curation to ensure the quality of the mapping. Relatedness among CUI-CUI pairs were largely extracted from the UMLS including several major classes such as *“may treat or may prevenf’, “classifies”, “differential diagnosis”, “method of’* and *“ca’Usatfoe”*. We additionally curated related codified feature pairs by mapping disorder CUis to PheCodes, drugs to the RxNorm, and procedures to CCS categories. These mapped code pairs are then further used to assess the ability to detect relatedness among codified features.

We evaluated different types of relationships by computing the cosine similarities be tween the embedding vectors of related pairs and randomly selected pairs. This allowed us to calculate the Area Under the Curve (AUC) of the cosine similarities, which measures the ability to distinguish known pairs from random pairs. The random pairs were chosen to have similar semantic characteristics as the related pairs. For instance, when examining the relationship “may treat or may prevent,” we only considered disease-drug pairs. We compared MultiReL embeddings to those from single EHR sources as well as the pre-trained language model embeddings. Since some EHR concept pairs may not appear in a single EHR system, we use the orthogonal translation of the EHR embeddings trained in the other system as pre-training (29) to expand the concept pool for a fair comparison. For pre-trained language models, we includedBioBert (30), PubmedBert (31), and SAPBert (32) represented by the descriptions of the code or CUI. These are variants of the Bidirectional Encoder Representations from Transformer (BERT) models fined tuned for biomedical text. For example, the SAPBert (Self-aligning pretrained BERT) leveraged the UMLS and PubmedBert pretrained with PubMed text while BioBert is pretrained on both general domain corpora and biomedical domain corpora. For CUis with different description terms, similar to how we train the LLM CODER embedding in ONCE, the pre-trained language embeddings are averaged across all terms that represent the CUI.

To further evaluate the quality of ONCE feature selection, we performed case studies for 8 diseases, namely coronary artery disease (CAD), rheumatoid arthritis (RA), Crohn’s disease (CD), ulcerative colitis (UC), heart failure (HF), type 1 diabetes (TlD), type 2 diabetes (T2D), and depressive disorder (Depression), for which we have manually curated gold standard labels, averaging 150 patients per disease on the true disease status from MGBB. To account for patients with no main ICD record and reflect the overall importance of features to a disease, we randomly sample from zero-count patients as part of the negative controls in the label data. For each of the 8 phenotypes, we generated ONCE features that included both codified and NLP CUI concepts. We assessed (i) the concordance between ONCE feature importance score and relevance score assigned by ChatGPT (33) and GPT-4 (34), as measured by rank correlation, for each disease, where the GPT-4/ChatGPT scores were obtained by prompting them to rate the relevance of the features and the given disease on a scale of O to 1 and (ii) the concordance between ONCE score, GPT-4 or ChatGPT feature score and the strength of association between the disease status and the presence of the concept quantified by the negative log_10_ p-value from Fisher’s exact test for the association with MGBB labeled data. The presence of the concept is defined as having at least 3 counts of the concept to ensure specificity. The more important a feature is for the disease, the smaller p-value one would expect from the association test. We thus use the observed association strength as a gold standard to assess the quality of the ONCE feature importance score. For the ranking-concordance evaluation, we included the union of features selected by ONCE, an equal number of top ranked features by GPT-4/ChatGPT, as well as a random sample of 100 EHR features as negative controls.

#### 2.3.2 Accuracy of KOMAP

We further validated the performance of KOMAP phenotyping using partially labeled EHR data from four healthcare systems: (i) Biobank cohort from MGB on 8 disease phenotypes including CAD, RA, CD, UC, HF, TlD, T2D, and Depression; (ii) University of Pittsburgh Medical Center (UPMC) on Alzheimer’s disease (AD) and multiple sclerosis (MS); (iii) VA on RA and HF; (iv) Boston Children’s Hospital on HF, Asthma, and CD. The sample sizes of labeled and unlabeled observations used for training and validating phenotyping algorithms for different diseases are provided in supplementary table 2.

All gold standard labels on the true disease status were curated via manual chart review, sampled from those with at least 1 PheCode of the condition, which is also the filter used to define the target population for the phenotyping algorithm. For all phenotypes, we use both the PheCode and CUI(s) corresponding to the disease as the main surrogates. Since there are two CUis for HF (C0018801 and C0018802) according to the search result from ONCE, we have 3 main surrogates for HF. We compare the KOMAP algorithm trained with both NLP and codified features to KOMAP algorithm trained with only codified features to evaluate the value of NLP concepts. As benchmarks, we additionally compared to the main PheCode (main code), the main NLP CUI feature (main CUI), as well as other weakly supervised algorithms, including PheNorm(12) and MAP(26), trained using patient level data. For the ONCE selected features, we further removed features that appear in fewer than 5% of patients.

We validated the performance of phenotyping algorithms against the gold standard labels for each phenotype. We report the average AUC of the algorithms across phenotypes within each institution. We further report the difference AUC between KOMAP algorithm trained with both codified and narrative features and other benchmark algorithms averaged across phenotypes within institution. We report the p-value for testing whether the AUC difference is zero based on a Z-test on the AUC difference with its standard error calculated via the bootstrap. To examine the performance of our proposed simulation based KOMAP accuracy evaluation, we compare the KOMAP algorithm accuracy calculated based on patient level data against via our simulation approach.

## 3 Results from Validation Studies

### 3.1 MultiReL Embeddings and ONCE Feature Selection

In Table 1, we present the average AUC in distinguishing similar or related pairs from random pairs of codified concepts, codified vs CUI concepts, and CUI concepts, based on pairwise cosine similarities generated from various embeddings. In general, MultiReL embeddings attained most robust performance in terms of the overall embedding quality in detecting both similarity and relatedness, compared to both embeddings trained mainly with a single EHR source or language models. EHR generated embeddings generally are substantially better in detecting relatedness due to their ability in capturing real world disease management patterns. MultiReL training is also substantially better at mitigating potential biases from a single EHR system. For example, for CUI-CUI pairs with nodes that only appear in one EHR system, MultiRel attained an average AUC of 0.929 for similarity and 0.820 for relatedness while the single EHR source training yielded a much lower AUC of 0.783 for similarity and 0.617 for relatedness. More detailed performance summary of the MultiReL embeddings compared with other embeddings is shown in supplementary tables 3 and 4.

**Table 1:** AUCs of between-vector cosine similarity in detecting known similar or related pairs of codified concepts (COD), NLP CUI concepts, and COD vs NLP CUI concepts for embeddings trained by MultiReL, MGB alone with VA pre-training, VA alone with MGB pre-training, BioBert, PubmedBert, and SAPBert. To further illustrate the advantage of multi-source learning, we also evaluate the accuracy in detecting relationship pairs that do not co-occur in at least one of the EHR data sources, denoted by subscript *8·*

|  | Type | MultiReL | MGB | VA | BioBert | PubmedBert | SAPBert |
| --- | --- | --- | --- | --- | --- | --- | --- |
| COD-COD | similar | **0.951** | 0.876 | 0.921 | 0.596 | 0.627 | 0.780 |
|  | similar<sub>s</sub> | **0.911** | 0.832 |  | 0.653 | 0.647 | 0.811 |
|  | related | **0.820** | 0.793 | 0.808 | 0.576 | 0.620 | 0.660 |
|  | related<sub>s</sub> | **0.819** | 0.802 |  | 0.671 | 0.689 | 0.725 |
| CUI-CUI | similar | **0.931** | 0.806 | 0.836 | 0.651 | 0.773 | 0.833 |
|  | similar<sub>s</sub> | **0.929** | 0.783 |  | 0.655 | 0.771 | 0.828 |
|  | related | **0.863** | 0.770 | 0.818 | 0.606 | 0.710 | 0.716 |
|  | related<sub>s</sub> | **0.820** | 0.617 |  | 0.495 | 0.687 | 0.700 |
| CUI-COD | similar | **0.947** | 0.878 | 0.906 | 0.560 | 0.623 | 0.809 |
|  | similar<sub>s</sub> | **0.922** | 0.841 |  | 0.496 | 0.423 | 0.688 |

#### 3.1.1 ONCE Feature Selection

To determine the ability of ONCE to identify relevant codified and narrative features for use in phenotyping, ranking scores were calculated to compare the ranking of ONCE-selected features to features that would be selected by large language model tools ChatGPT and GPT4.

The ONCE feature importance scores largely agree with importance scores assigned by GPT4 and chatGPT as shown in figure 3. The average concordance was 0.638 between ONCE importance score and GPT4 importance score ranged and 0.612 for narrative features. ChatGPT shares a slightly higher similarity with ONCE when assigning the relevance score, given an average concordance of 0.657 for codified features and 0.649 for narrative features. As a benchmark, the average concordance between GPT-4 and ChatGPT is 0.782 for codified features and 0.787 for narrative features. When compared against observed association strengths from labeled EHR data, ONCE importance scores generally attained a higher concordance compared to those from GPT4 or chatGPT as shown in Figure 4. The average concordance was 0.412, 0.360, and 0.346 for ONCE, GPT4 and ChatGPT ranking of codified features; 0.527, 0.407 and 0.434 for narrative features.

**Figure 3:**
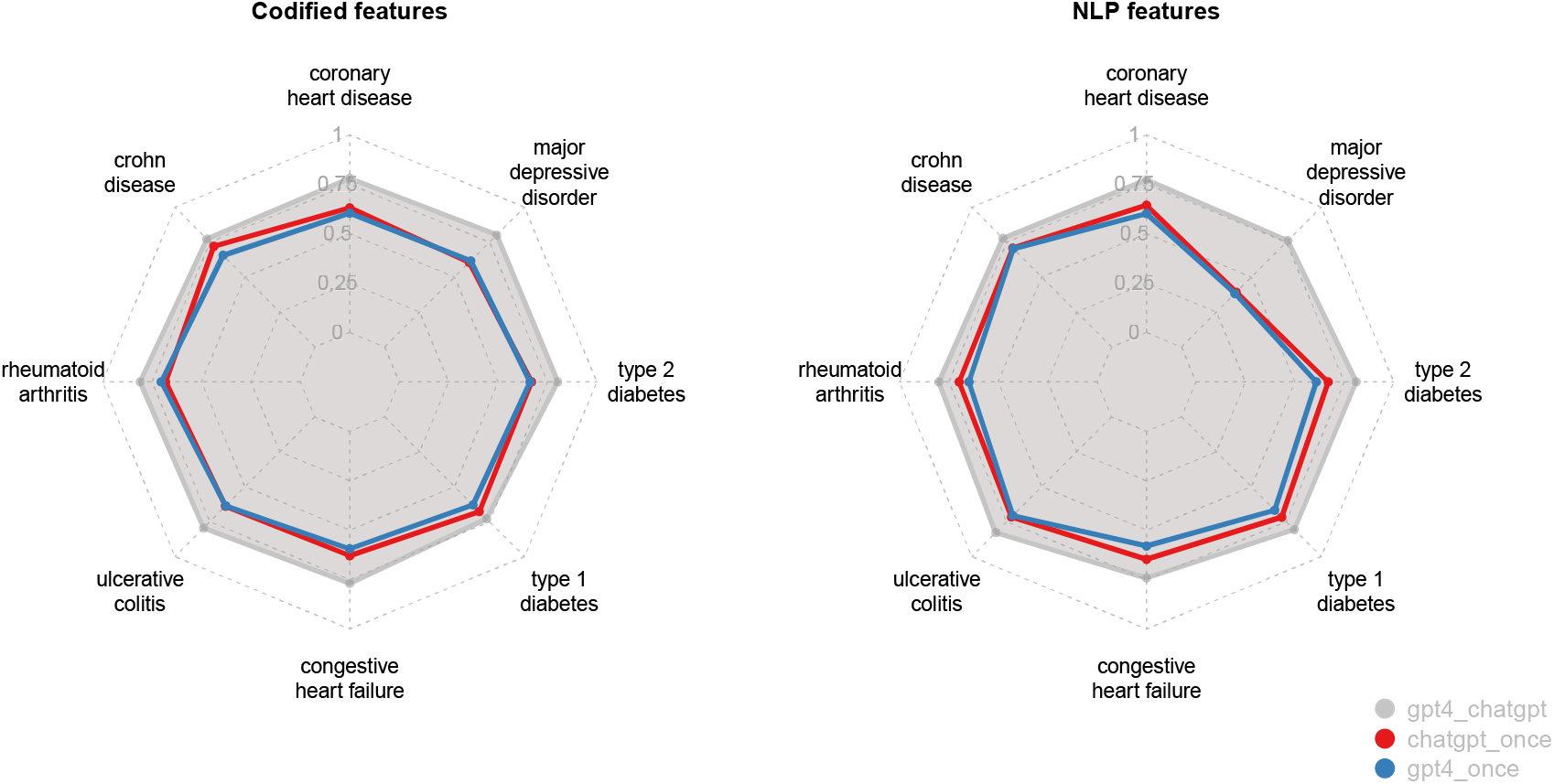
Disease-specific Kendall’s ranking correlation between pairs of ONCE, ChatGPT and GPT4 feature scores for 8 diseases.

**Figure 4:**
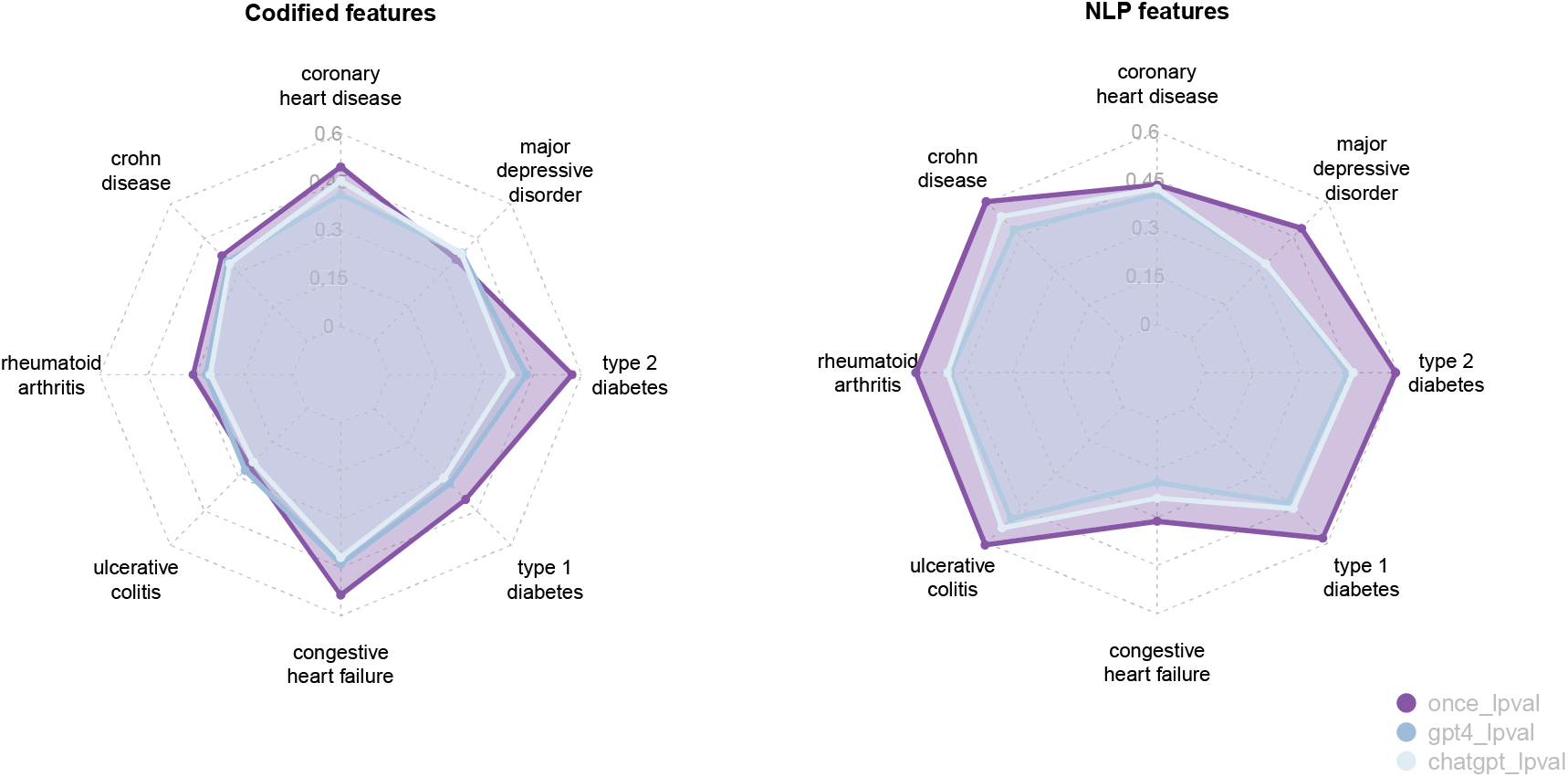
Disease-specific Kendall’s ranking correlation between the negative log_10_ pvalue from Fisher’s exact test for label data in MGB and ONCE/ChatGPT /GPT4 scores. For each subject and each feature, Fisher’s exact test classifies patients by its labels as well as whether it has at least 3 counts of the feature.

### 3.2 KOMAP Phenotyping

Figure 5 shows the average AUC of phenotyping algorithms within each of the 4 institutions trained by KOMAP, PheNorm, MAP, main code, main CUI and supervised methods. Taking MGB site as an example, averaging over the 8 diseases, the KOMAP algorithm trained with both codified and NLP data achieved accuracy (AUC: 0.956, SE: 0.006) that is comparable to that of PheNorm (AUC: 0.959, sd: 0.007), and significantly higher than that of supervised learning model with both codified and NLP data(AUC difference: 0.036, p-value: 8.76e-05), MAP (AUC difference: 0.030, p-value: 7.82e-05), main ICD (AUC difference: 0.065, p-value: 2.44e-06), and main CUI (AUC difference: 0.057, p-value: l.29e-06). Similar patterns are observed when comparing the algorithms trained with codified data only. Due to the small number of labels, supervised algorithms generally suffered from overfitting and attained low accuracy. Including NLP surrogate as well as narrative features can substantially improve the phenotyping accuracy. Comparing KOMAP trained with NLP plus codified versus codified alone, the average gain in AUC was 0.073 (p-value: 0.014) at BCH, 0.051 (p-value: 0.008) at VA, 0.027 (p-value: < 0.001) at MGB, and 0.052 (p-value: < 0.001) at UPMC. For individual phenotypes, the difference is the most striking for depression at MGB (KOMAP both AUC: 0.943; KOMAP codified AUC: 0.870; p-value for AUC difference: 0.027).

**Figure 5:**
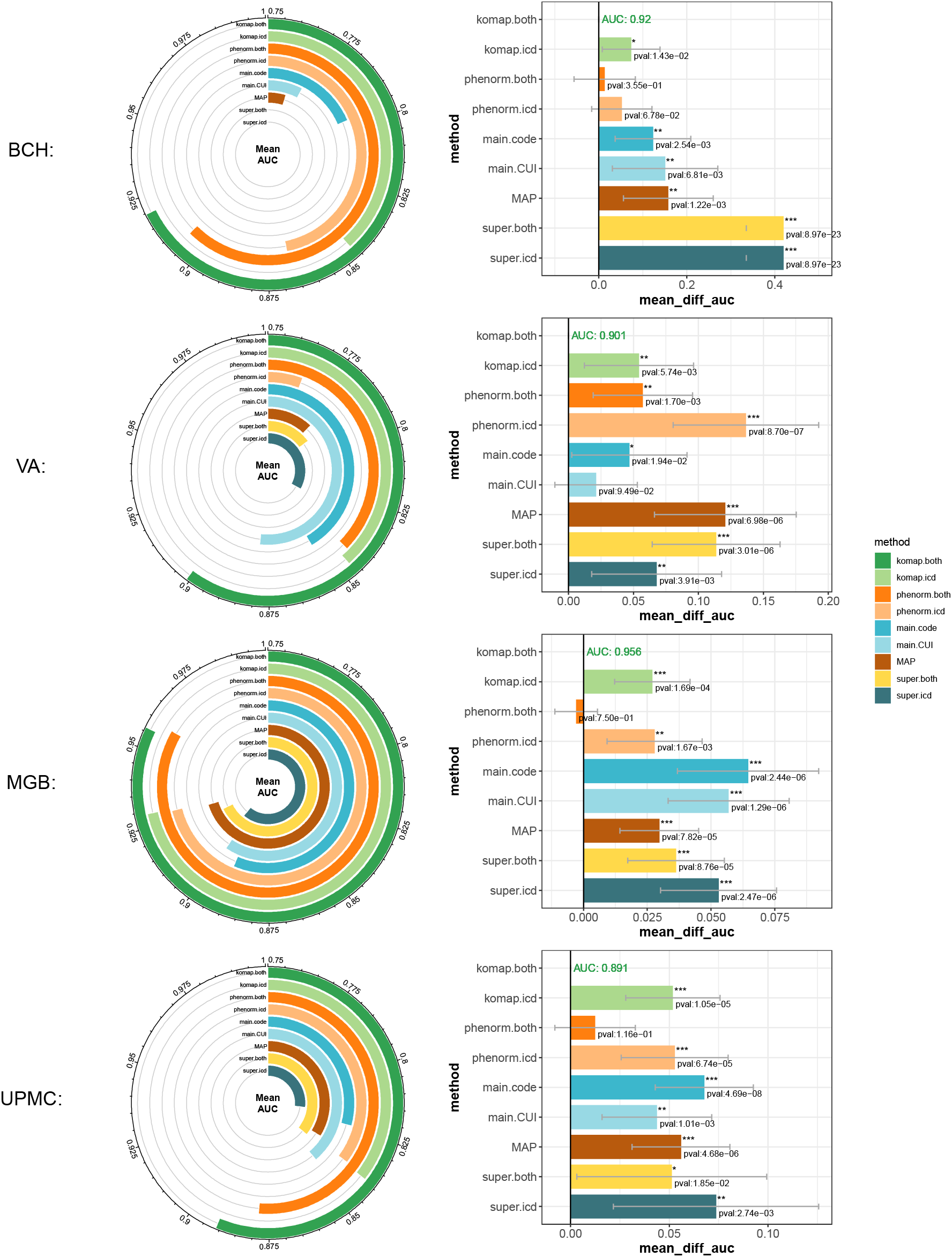
Average AUC (left) and average AUC difference (right) over phenotypes against KOMAP with both codified data and CUis across sites. Competitors include PheNorm with both codified data and CUis; supervised model with both codified data and CUis:1: 0MAP with only codified data; PheNorm with only codified data; supervised model with only codified data; main ICD; main CUI and MAP.

Although KOMAP and PheNorm attain similar performance, KOMAP algorithms are generally more sparse with non-zero weights assigned to fewer features. To illustrate the superiority of KOMAP, we also visualize the coefficients of features for depression estimated by KOMAP, PheNorm and supervised-learning model with codified (and NLP data) to see the difference. In figure 6, coefficients estimated from KOMAP behaved like a combination of those from the supervised model and PheNorm. Meaningful features that were assigned high importance in the supervised-learning model such as venlafaxine, bupropion and hydroxyzine also shared non-zero effect from our KOMAP algorithm. Besides, KOMAP picked up some features that were only non-zero from PheNorm such as depressed mood, major depressive disorder and mirtazapine. Coefficients for other diseases have been shown in the appendix.

**Figure 6:**
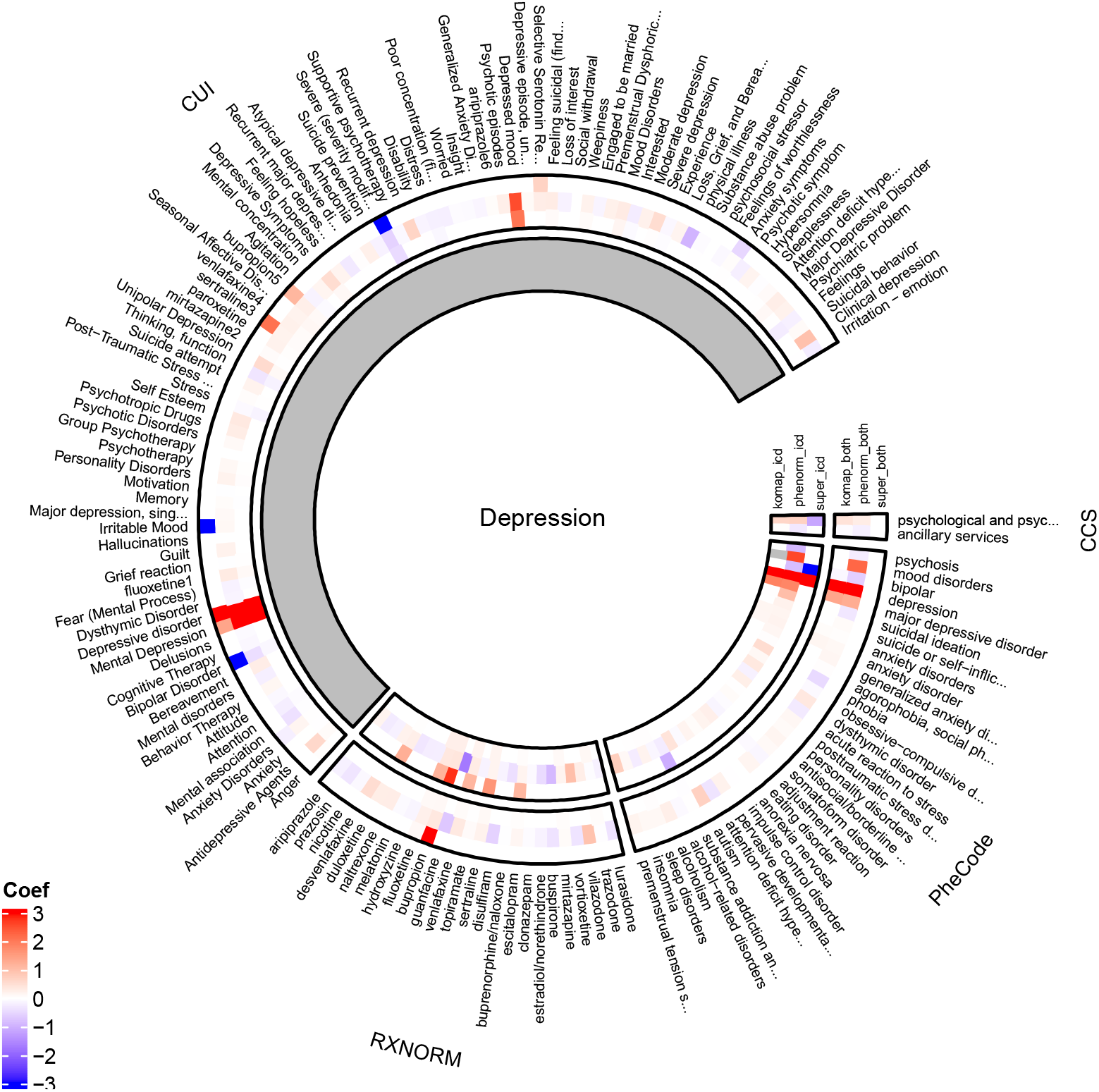
Coefficients of features for depression estimated by KOMAP, PheNorm and supervised-learning model with codified and NLP data (outer circles) or only with codified data (inner circles).

We also visualized our proposed simulated AUC versus the real AUC for KOMAP with only codified features and with both codified and narrative features in figure 7. The average absolute difference between the simulated and the real AUC among 8 diseases in MGB site is 0.016 for codified only KOMAP and 0.008 for KOMAP with codified and narrative features, which is relatively small. In 7 out of 8 diseases, KOMAP with codified and narrative features tends to slightly overestimate the AUC while for depression, simulated AUC generated from both versions of KOMAP underestimated the result.

**Figure 7:**
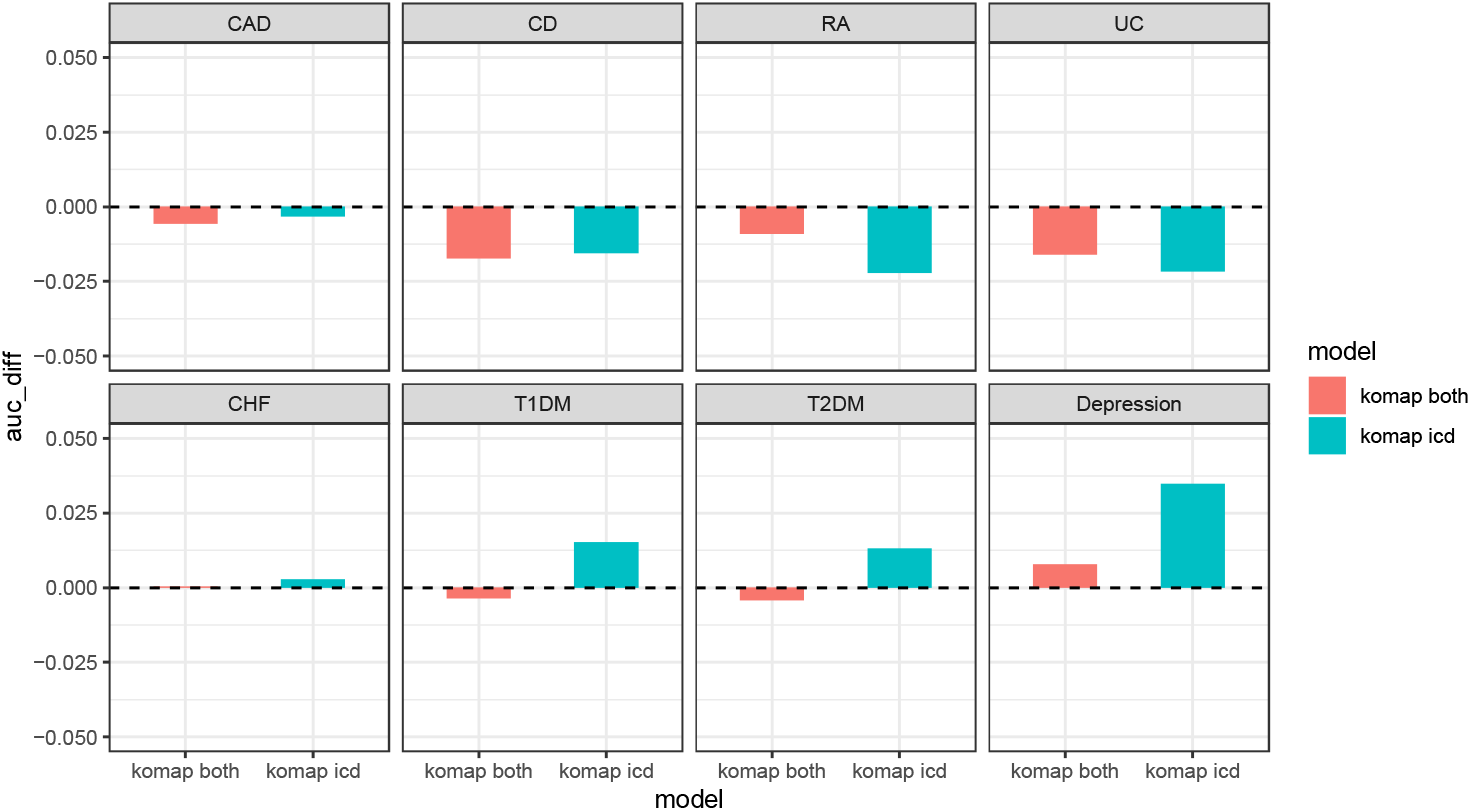
Difference between the real AUC and the simulated AUC for the 8 MGB phenotypes generated from KOMAP with both codified data and CUis and KOMAP with only codified data.

### 3.3 Two Web APis

We implemented two R-shiny powered online web-APis (https://shiny.parse-health.org/ONCE/ and https://shiny.parse-health.org/KOMAP/) to realize the functionality of the ONCE system and KOMAP algorithm.

## 4 Discussion

In this paper, we have decomposed the EHR-driven phenotyping task into automated feature selection and algorithm training without gold-standard labels. The former goal has been achieved by ONCE, and the latter goal has now been achieved by KOMAP. The utilization of the ONCE feature selection tool and KOMAP training algorithm offers a significant boost to researchers’ capacity to incorporate previous knowledge regarding EHR features and to reduce n01se in EHR data. This, in turn, leads to improvements in the reliability and applicability of subsequent predictive modeling.

Specifically, the evaluation results of ONCE feature selection against GPT4/chatGPT indicate that the feature importance score generated by ONCE not only agrees largely with the biomedical knowledge that currently exists in the state-of-art AI models, but also matches the feature-disease relevance to a large extent. Even though for certain diseases, there exists inconsistency between ONCE feature scores and AI-generated scores (e.g., narrative features for *major depressfoe disorder)*, the higher concordance between the Fisher test significance and ONCE score (ONCE: 0.485; GPT-4: 0.329; ChatGPT: 0.328) further illustrates the superiority of ONCE in identifying more relevant EHR concepts. As more institutions generate co-occurrence matrices of their EHR features and large language models get updated over time, our MultiReL algorithm can also be updated with these new data to further improve the quality of the embeddings and generate a more comprehensive list of disease-related features.

Benefiting from the ONCE feature selection result, KOMAP has unique advantages with regard to other phenotyping algorithms. In terms of computation time, KOMAP improves its speed greatly due to the pre-selection step conducted by ONCE that screens out unrelated features. The algorithm is further accelerated by conducting regression on the feature space (ℝ^*p*×*p*^) instead of the original individual data space (ℝ^*n*×*p*^) without sacrificing the accuracy. Compared to other established models based on a humongous volume of patient-level data, KOMAP only requires a few seconds to complete the phenotyping as well as the validation task, which renders it a highly scalable algorithm.

Another plus of KOMAP is its capacity of dealing with multiple surrogates for one disease. The most common case is when the codified data and NLP data are all at hand, the current KOMAP algorithm allows users to specify both the main PheCode and the main CUI corresponding to the disease of interest as surrogates. In general, across 4 institutions, training KOMAP with the additional main CUI (and also related NLP concepts) improved phenotyping accuracy. Such an improvement persists even if the main CUI counts suffer from worse performance compared to the single main ICD feature counts. For instance, the average AUC of the main ICD counts in BCH is 0.797 while that of the main CUI counts is only 0.769. However, by integrating both surrogates, the performance of KOMAP rises from 0.846 trained with the main ICD surrogate and codified features to 0.898 trained with both surrogates and codified and narrative features. It could also occur when there is no single mapping between a disease and its NLP / codified feature. For example, as suggested by the search result from ONCE, heart disease may have multiple narrative features (e.g., C0018801 and C0018802) that can serve as the main surrogate. In order to harness all the useful information in multiple surrogate candidates, KOMAP is equipped with the principle component analysis step. It can merge disease scores derived from different surrogates into one, and the same applies to the feature coefficients, which eventually rendered a higher and more stable phenotyping accuracy compared to the single surrogate case, as shown in figure 8. In order to compare across sites, AUC on the figure has been standardized among different surrogate combinations. Notice that treating C0018801 as the single surrogate works well for BCH (0.827, 2nd highest) and MGB (0.957, 2nd highest) data, but poorly for VA patients (0.827, lowest). On the other hand, combining C0018802 and PheCode:428 as two main surrogates achieves the highest AUC (0.878) for VA data. Still, the three-surrogate version of KOMAP (BCH AUC: 0.84; VA AUC: 0.873; MGB AUC: 0.962) is strongly recommended given its robustness regardless of the heterogeneous performance of different single surrogates. The superiority of KOMAP is also shown by the comparable or even higher accuracy with only summary-level (i.e., covariance matrices) data as input. Existing phenotyping pipelines largely require training with patient-level data with gold labels while the proposed KOMAP algorithm removes the need for manual annotation which is resource intensive and not scal able. In addition, KOMAP simulation-driven validation algorithm, which is also based on summary-level data, well approximates the true accuracy obtained based on the individual level data across all the studied phenotypes. However, one potential limitation exists since KOMAP would require approximating the disease-specific covariance matrix using patients whose count of main PheCode is larger than 0. The estimate could be quite unstable if the PheCode suffers from a very low prevalence rate (e.g., some rare 1-digit or 2-digit PheCode). One possible solution is to expand the population to include patients with at least one rolled up integer-level PheCode, instead of just the main PheCode, then perform KOMAP on the higher-level covariance matrix. By doing so, we not only achieve a larger sample size but also obtain richer information brought by patients with other sibling/parent/child PheCodes. This may help to phenotype the target disease due to the shared characteristics as well as distinctive features among different PheCodes under the same integer category. Specifically, we have noticed that by merging TlD (PheCode:250.1) and T2D (PheCode:250.2) patients together, the prediction performance of our KOMAP improved to a certain extent (TlD with codified data only: from 0.922 to 0.933; TlD with codified and NLP data: from 0.955 to 0.967). Though performance for T2D deteriorated slightly (codified data only: from 0.918 to 0.917; codified and NLP data: from 0.932 to 0.928). Similar improvements occurred if we applied the combined covariance matrix to CD (PheCode:555.1) and UC (PheCode:555.2) patients (UC with codified data only: from 0.933 to 0.950; UC with codified and NLP data: from 0.954 to 0.964; CD with codified data only: from 0.942 to 0.940; CD with codified and NLP data: from 0.962 to 0.963;). Details have been shown in figure 9.

**Figure 8:**
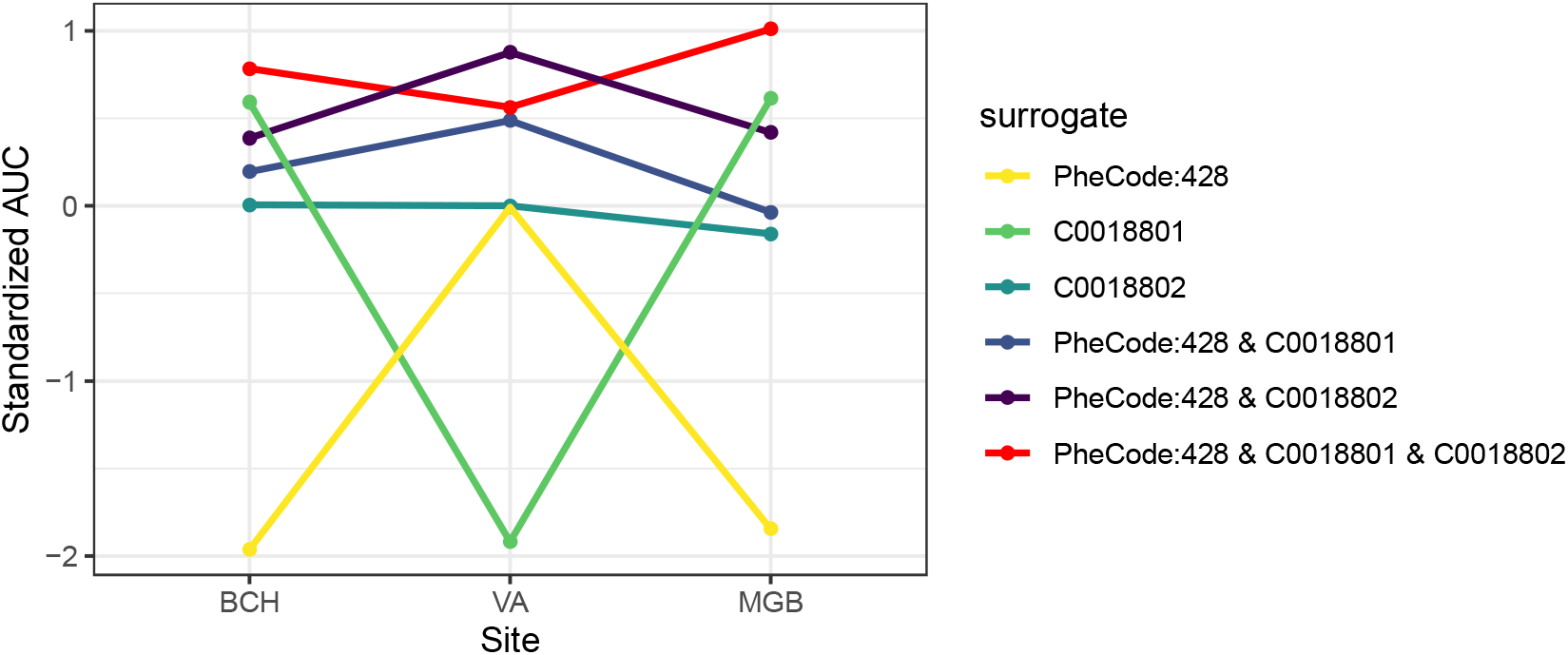
Standardized heart failure AUC ofKOMAP across BCH, MGB and VA with different combinations of surrogates. Possible choices include the main ICD PheCode:428 and two important CUis – C0018801 and C0018802.

**Figure 9:**
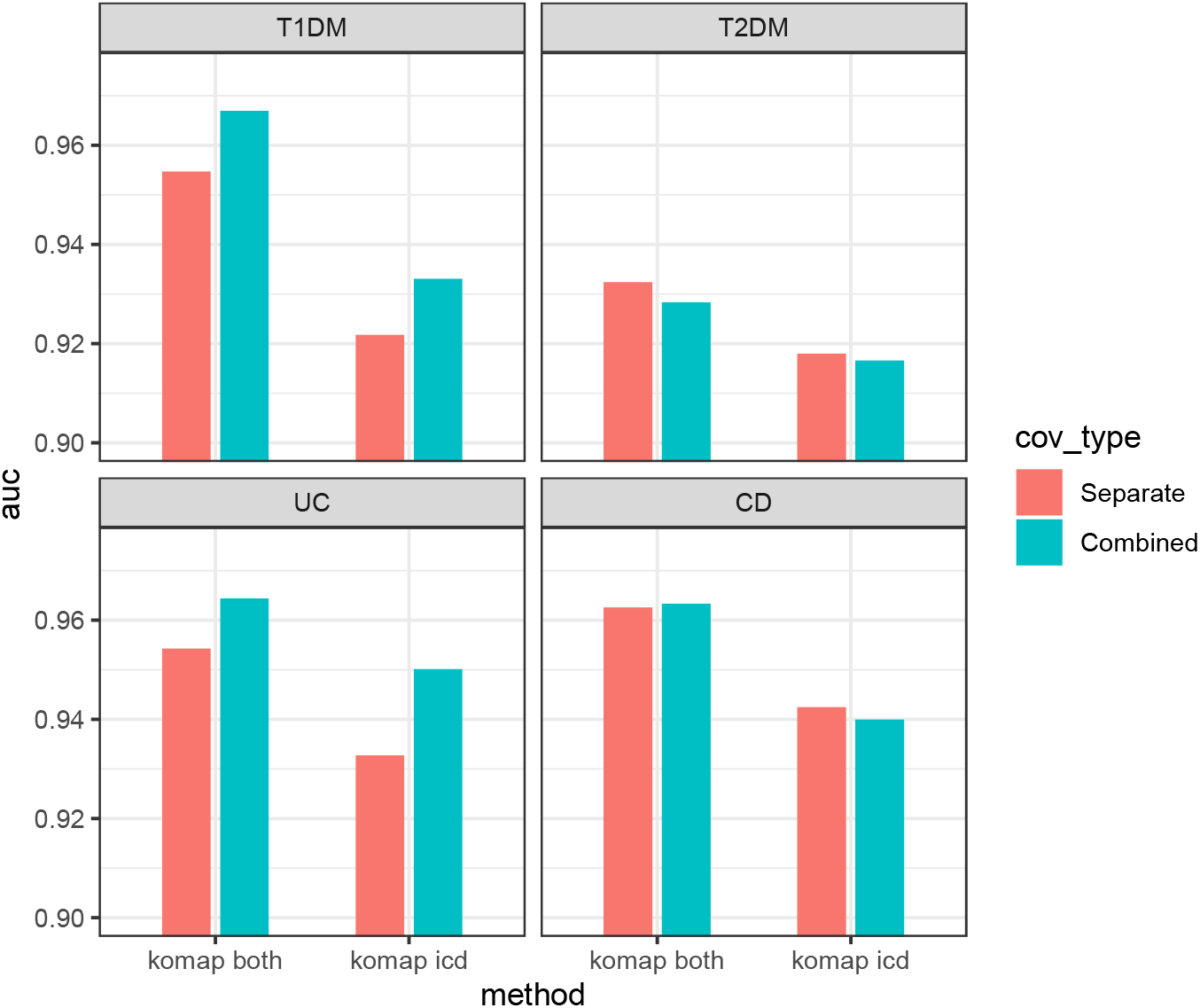
Upper two panels: AUC for TlD and T2D generated from the combined/separated version of KOMAP with both codified and NLP data or with only codified data; lower two panels: AUC for UC and CD generated from the combined/separated version of KOMAP with both codified and NLP data or with only codified data.

In fact, both the MultiReL representation learning algorithm and the KOMAP pheno typing algorithm operate effectively with summarized data, enabling collaborative learning opportunities among multiple institutions. Yet, data from different institutions might be heterogeneous due to different practices and coding or writing habits of clinical professionals at different institutions. These differences result in data heterogeneity in not only codified data but also data from narrative notes. Many federated learning methods have developed in past decades, such as Federated stochastic gradient descent (FedSGD) (35) Federated averaging algorithm (FedAVG) (35), and Cyclical weight transfer (CWT) (36). Co-training a phenotyping algorithm by allowing researchers from different institutions to upload summary level data to our Webapp and accounting data heterogeneity from different institutions is one of our future steps to improve the robustness of our online phenotyping pipeline.

## Data Availability

Summary data used for illustration of the method described in the manuscript is available online as part of a web app at https://shiny.parse-health.org/KOMAP/

https://shiny.parse-health.org/ONCE/

## Acknowledgements

This project was supported by NIH grants 1OT2OD032581, R0l HL089778 and R0l LM013614, P30 AR072577, and the Million Veteran Program, Department of Veterans Affairs, Office of Research and Development, Veterans Health Administration, and was supported by the award #MVP000. This research used resources from the Knowledge Discovery Infrastructure at Oak Ridge National Laboratory, which is supported by the Office of Science of the US Department of Energy under Contract No. DE-AC05-00OR22725. This publication does not represent the views of the Department of Veterans Affairs or the U.S. government. This project was also supported by NCATS U0l TR002623 and by the PrecisionLink Biobank at Boston Children’s Hospital.

## Author Contributions

XX: Methodology, software, writing. SMS: Data curation, software, writing. ML: Method ology, software, writing. CH: Data curation. CLB: Data curation. VAP: Data curation. DZ: Methodology, software. LW: Data curation. ER: Resources. LC: Data curation. YLH: Data curation. AG: Data curation. KDM: Data curation. SC: Data curation. ZX: Data curation. KC: Data curation. JMG: Data curation. KPL: Data curation. TC: Conceptualization, supervision, methodology, writing. TC: Conceptualization, supervision, writing.

